# Prevention of canine visceral leishmaniasis in Brazil: a prospective study for evaluating the impact and operational difficulties of mass use of deltamethrin impregnated collars in Minas Gerais

**DOI:** 10.1101/2025.10.31.25339244

**Authors:** Lucas Edel Donato, Marilia Rocha Fonseca, Guilherme Loureiro Werneck, Fredy Galvis Ovallos

## Abstract

**Background:** Canine visceral leishmaniasis (CVL) remains a major public health challenge in Brazil, where domestic dogs are primary urban reservoirs of *Leishmania infantum*. Traditional control strategies, such as euthanasia of seropositive dogs, face operational limitations and ethical concerns. Insecticide-impregnated collars, particularly those containing 4% deltamethrin (DMC), have emerged as a promising alternative for reducing transmission.

**Methodology/Principal Findings:** A prospective cohort study was conducted in Montes Claros, Minas Gerais, from August 2021 to August 2023, involving 296 dogs divided into two groups: DMC and DMC plus reinforcement device (DMC+RD). Follow-up evaluations occurred every six months over four cycles. The DMC+RD group showed a 41% reduced chance of infection (OR = 0.59; 95% CI: 0.27–1.31) compared to the DMC group. Loss of follow-up was frequent, with 38% overall loss after 24 months, and 50% of owners unable to identify when the loss occurred. Although no statistically significant differences in collar retention were observed, the reinforced device showed a trend toward improved collar permanence.

**Conclusions/Significance:** High effectiveness of DMC applied in a mass population control program was observed, as well that the improvement of the collar design could contribute to reducing the loss of collars and increase the protection. Nevertheless, other factors related to the owners’ engagement, operational difficulties, and dog population dynamic, can influence the outcomes of the implementation strategy. These results contribute to optimizing CVL control programs and support the scalability of collar-based interventions in endemic regions.

**Author Summary:** In Brazil, canine visceral leishmaniasis (CVL) remains a major public health challenge due to its role in maintaining the transmission of *Leishmania infantum*, the parasite responsible for visceral leishmaniasis in humans. To address this, our study evaluated the effectiveness of collars impregnated with 4% deltamethrin in preventing infection in dogs, and tested a reinforcement device designed to reduce collar loss. We followed a cohort of dogs over two years in an endemic urban area, comparing those using the standard collar with those using the reinforced version. Our findings showed that both collar types helped reduce infection rates, but dogs with the reinforced collar had a lower chance of losing it, although the difference was not statistically significant. We also observed that many dog owners were unaware of when collars were lost, highlighting the need for better community engagement. This study provides valuable insights into the operational challenges of implementing this strategy at scale and suggests that improving collar design and owner participation could enhance the success of CVL prevention programs.

## Introduction

Leishmaniasis is an infectious, tropical, and neglected disease caused by protozoa of the genus *Leishmania*. Among the known clinical forms, visceral leishmaniasis (VL) is the most serious and can lead to death if not treated properly [1]. In the Americas, the main species involved is *Leishmania infantum*, which is transmitted predominantly by the bite of the female sand fly *Lutzomyia longipalpis* [2, 3]. Domestic dogs are recognized as the main urban reservoirs of the parasite, playing a central role in maintaining the transmission cycle [4]. Canine visceral leishmaniasis (CVL), therefore, represents not only an animal health problem but also a challenge for public health, being equally classified as a neglected disease with limited options for effective control [5].

In Brazil, the CVL control strategies face operational and epidemiological obstacles, especially in urban settings. The euthanasia of seropositive dogs, although widely adopted, lacks robust evidence regarding its effectiveness in reducing substantially the transmission [6, 7]. This limitation may be related to rapid population replacement by young dogs that are more susceptible to infection and the low accuracy of serological tests, which can result in both the elimination of non-infectious animals and the permanence of highly infectious dogs [8]. Additionally, the VL spatial distribution in Brazil shows differences in its occurrence, e suggesting structural inequities associated to its occurrence [9, 10]

Given these difficulties and the need for more effective and acceptable strategies, the use of insecticide-impregnated collars has emerged as a promising approach to preventing canine infection [11]. These products act through a mechanism of action that aims to prevent contact between the vector and the host, promoting a repellent and/or insecticidal effect. Thus, they reduce the probability of bites and, consequently, the transmission of the parasite [12]. In Brazil, the effectiveness of collars impregnated with 4% deltamethrin (DMC) has been evaluated since the early 2000s, with studies demonstrating a significant impact on reducing infection in dogs [13, 14] and, more recently, in humans [15].

In a cohort study conducted in an endemic area, a 63% reduction in the incidence of canine infection was observed among animals that wore the collar, compared to those in the control area [16]. In addition, a community intervention study evaluating the impact of the same intervention identified a 27% reduction in the incidence of human cases in areas where collars were distributed, compared to areas without the intervention [15].

The operationalization of a population intervention based on DMC faces challenges inherent to the application of any strategy in the field, with the loss of collars between collaring cycles being one of the main specific obstacles to its effectiveness. In the municipality of Monte Gordo-BA, for example, losses of up to 64.7% were recorded, with less than 42% of dogs keeping their collars until the subsequent evaluation, which impair the continuous protective coverage and might impact the effectiveness of the intervention [17]. In contrast, a community trial conducted in Iran showed significantly lower losses, with an annual average of only 6.9% and rapid replacement in more than 90% of cases, maintaining an average coverage of 87% of dogs [18]. These findings show that the effectiveness of the strategy depends not only on the efficacy of the product, but also on factors related to the implementation, including community adherence.

Despite advances in the use of DMC as a VL control strategy, important gaps in knowledge remain that limit the optimization of this intervention. Few studies have evaluated the impact of complementary devices or strategies to prevent collar loss, a factor that can significantly impair coverage and continuous protection of dogs. Furthermore, it is not sufficiently clear how long dogs remain effectively collared in real field conditions, nor how this duration influences the effectiveness of the intervention. Although the study by [18] demonstrated a relatively low loss rate with rapid replacement, the variability between operational contexts in different communities suggests the need to evaluate aspects that can affect the intervention and identify opportunities to improve the sustainability of the strategy in public health programs aimed at controlling visceral leishmaniasis. In Brazil, the use of DMC for the prevention of VL was incorporated in 2021 to be applied in priority areas defined by the Brazilian Ministry of Health’s Visceral Leishmaniasis Surveillance and Control Program (VLSCP) [19]. Therefore, the present study aims to evaluate aspects related to the mass use of DMC in the canine population under real operational conditions and identify potential opportunities to improve its implementations.

## Methods

### Type and location of study

This was a prospective cohort study including dog population undergoing intervention with DM collars, conducted between August 2021 and August 2023. The reinforcement device tested was a nylon collar to which the DMC collar was attached (Fig 1A, Fig 1B, Fig 2A, Fig 2B). The outcomes evaluated were the effectiveness of the collar protection for the infection with *Le. infantum* and the effectiveness of a reinforcement device on the reduction of collar loss. The study was conducted in the municipality of Montes Claros, located in the north of Minas Gerais state, about 422 km from the capital. It occupies an area of 3,589,811 km and has a population of 422,793 inhabitants, ranking as the fifth most populous municipality in the state (20). This municipality was selected according to the following criteria: 1) It is an endemic area for VL; 2) it is classified as a very intense transmission area; 3) The mass use of the DMC collar being implemented at the start of the study; and 4) the availability of human resources to undertake the field activities and local health managers consent.

**Fig 1.**
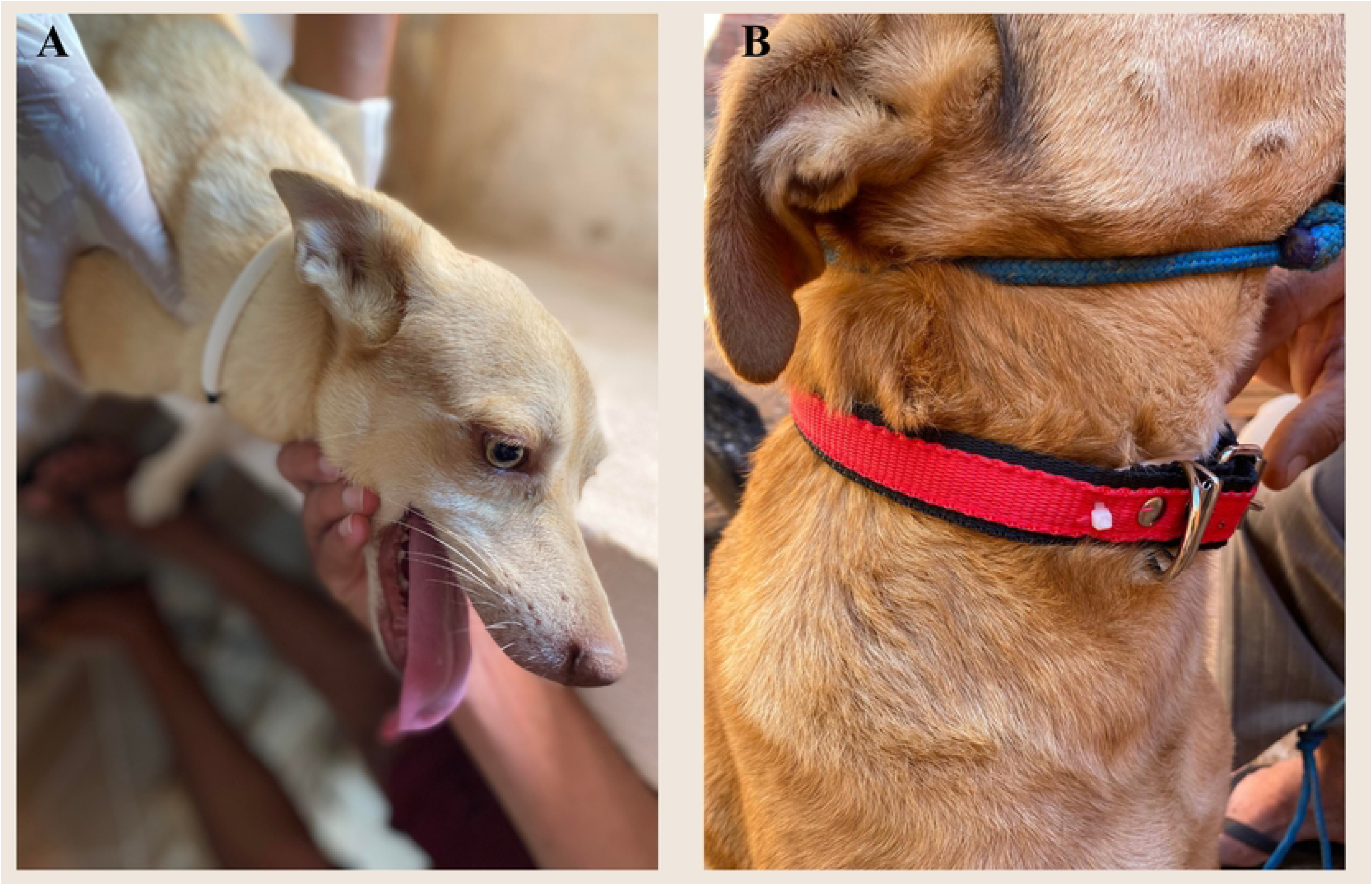
Insecticide DMC collar: conventional use and structural reinforcement with nylon. (A) External view showing the nylon reinforcement device coupled to the insecticide-impregnated DMC collar. (B) Internal view highlighting the insecticide matrix embedded within the collar structure.

**Fig 2.**
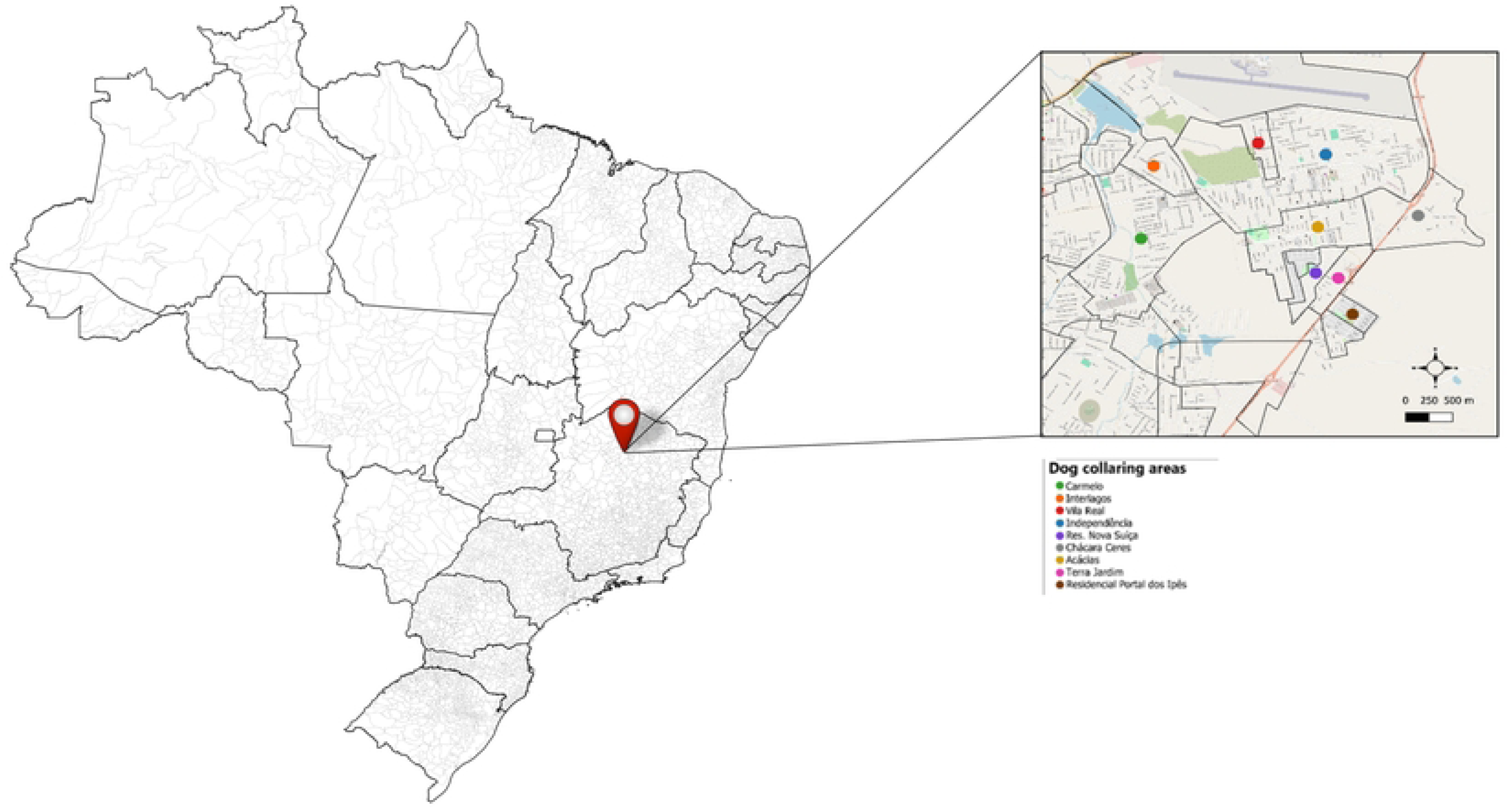
Designated Zones for Canine Insecticide Collar Deployment in Montes Claros Municipality.

### Study design

The area under intervention was initially georeferenced using the software QGIS 3.34 [21]. After that, ten blocks were randomly selected to undertake the dog recruitment.

These areas, called Local Work Areas (LWA), had homogeneous characteristics, considering the incidence in humans, canine density and prevalence, and human development index. After selecting the blocks, all households in each LWA were visited, and in those with dog presence, the owners were invited to participate in the study (Figure 2).

The dog population included in the study was divided into two groups: deltamethrin collar (DMC) and deltamethrin collar plus reinforcement device (DMC+RD). The randomization was performed at the block level, with each block being randomly assigned to receive either the intervention (DMC+RD) or the control (DMC) (Fig 3A, Fig 3B).

**Fig 3.**
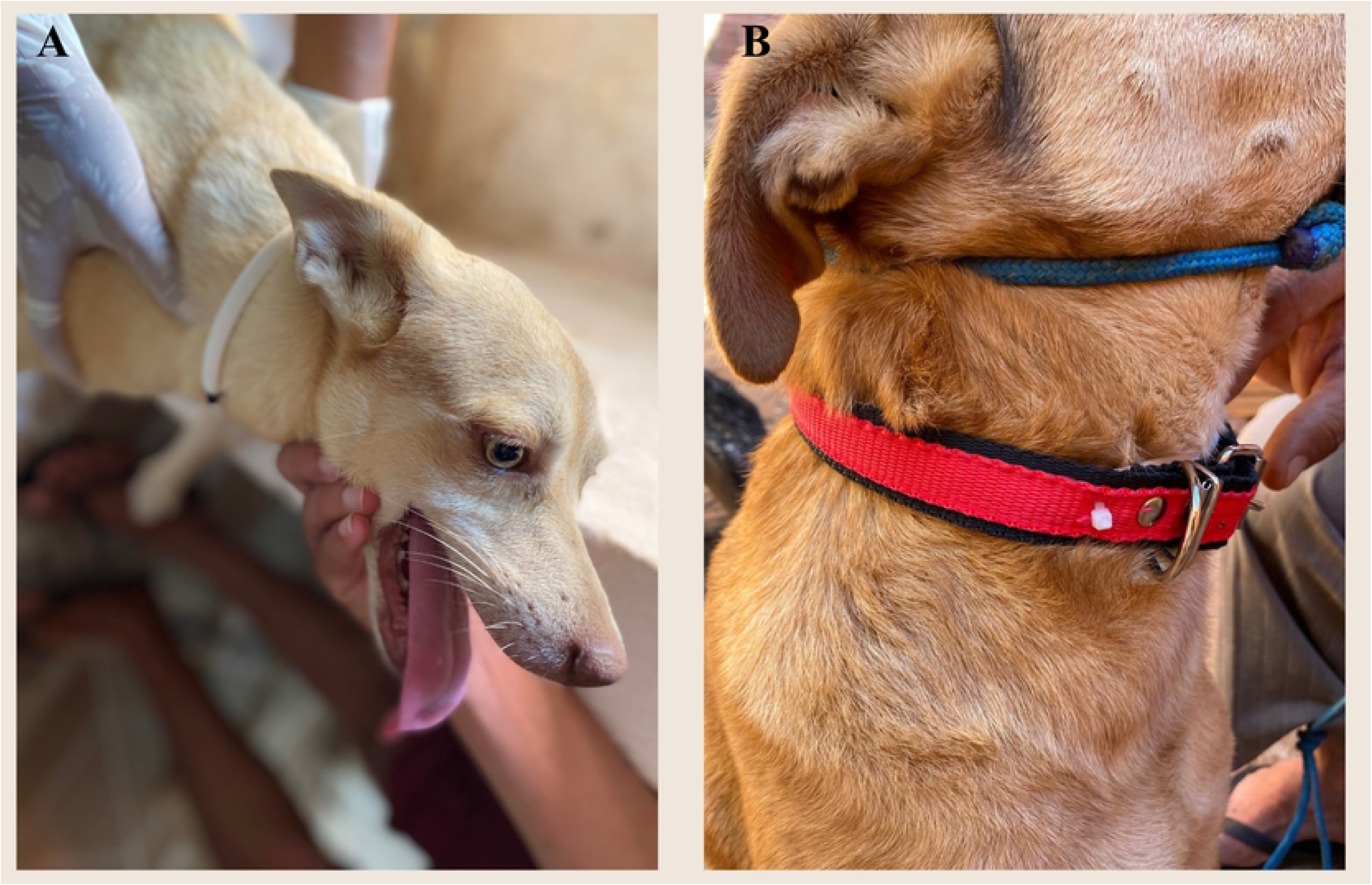
Use of insecticide-impregnated collar in dogs: standard configuration and reinforced version with nylon device. (A) Dog wearing standard DMC collar. (B) Deltamethrin collar plus reinforcement device (DMC+RD)

### Inclusion/exclusion criteria

Owned dogs, living in the domicile, independent of the age, with the guardian’s consent for using the DMC, blood sample collection and response to a questionnaire on variables associated with dogs and domicile characteristics were included. Animals seroreagent for infection with *Le. infantum* were excluded from follow-up.

### Statistical power and sample size

The average prevalence of infection/disease in the municipality was 7%, and the canine population in the intervention area was estimated at 700 dogs, according to information obtained from the canine census conducted by the Zoonosis Surveillance Unit. With the 296 recruited dogs at baseline (150 in the DMC group and 146 in the DMC+RD group) the study had an 80% statistical power to detect a relative risk (RR) of infection equal orf higher than three, with a 95% confidence level.

### Cohort follow-up

The follow-up was conducted at four evaluation times (T), designated as T0 (baseline), T1, T2, and T3, with a six-month interval between each evaluation. At each time, venous blood was collected to evaluate the infection status. Animals diagnosed as reagents were excluded from the cohort, and the collars were replaced according to the group.

A team of endemic disease control agents, linked to the municipality’s Zoonosis Surveillance Unit, was assigned to carry out the field activities. After specific training, the team conducted serological surveys on the dogs, collaring, and standardized data collection using a previously validated questionnaire. This instrument contained information related to the guardian, individual characteristics of the animals, as well as data on serological diagnoses and collaring, clinical status of the dog, presence or absence of collars, and the dogs’ usual sleeping place.

The collection of biological samples followed the methodology adopted in serological surveys as recommended by the VLSCP. It was performed by puncturing the brachycephalic vein and, when necessary, the jugular vein. The minimum amount determined was 3 mL, transferred to a tube without an anticoagulant. The samples were centrifuged for 10 minutes at a speed of 2,000-3,000 revolutions per minute (rpm). To comply with the recommendations of the Brazilian Ministry of Health, a serial methodology was used for the diagnosis of CVL, involving a screening test followed by a confirmatory test. For screening, the immunochromatography technique was employed using the Dual Path Platform rapid test (Bio-Manguinhos laboratory), with serum following the manufacturer’s instructions. Serum samples from reagent dogs in this test were tested using Enzyme-Linked Immunosorbent Assay ELISA (Bio-Manguinhos laboratory). This step was undertaken at the Ezequiel Dias Foundation in Belo Horizonte, Minas Gerais. Dogs were considered positive for CVL when reagent in both tests.

### Statistical analysis

A descriptive analysis was performed on the dogs’ characteristics, including variables such as age, sex, breed, coat type, sleeping location, and housing status. Categorical variables were presented by absolute and relative frequencies. To evaluate the association between the *Le. infantum* infection and the type of collar used (DMC or DMC+RD), a binary logistic regression analysis was performed. The dependent variable was infection status (infected vs. uninfected), and the main predictor variable was the type of collar. The odds ratio (OR) of infection was estimated, with a 95% confidence interval.

The longitudinal evaluation of serological status was conducted considering the different follow-up times (T). For this purpose, survival analysis (time to seroconversion or exclusion from the cohort) was applied, using Kaplan-Meier curves to estimate the probability of dogs remaining non-reagent over time. The comparison between groups was performed using the Log-Rank test.

Regarding the probability of collar loss (outcome) between the comparison groups (DMC or DMC+RD), a binary logistic regression was used. For each observation time (T1, T2, and T3), a model was adjusted with the binary dependent variable and the categorical independent variable (group: DMC+RD vs. DMC). All analyses were performed using Jamovi and Epi Info. The level of significance adopted was 5% (p < 0.05).

### Ethical aspects

This study was submitted to the Animal Ethics Committee of the Oswaldo Cruz Foundation and approved under protocol number LW-18/21. The research, including a questionnaire aimed at characterizing the canine population, was submitted to the Research Ethics Committee of the State University of Montes Claros and approved under opinion number 5.408.691.

## RESULTS

### Dog population characterization

A total of 296 dogs were recruited in the study, being 56.7% (n = 168) female, with an average age between 1 and 3 years. There was an equivalence between small and medium-sized dogs, approximately 38% for each. Regarding coat type, there was a higher frequency of short-haired dogs, representing 59.8% (n = 177). The description of the dog population studied is presented in Table 1.

Most dogs (89.9%, n = 266/296) were owned and lived within household premises. It was also observed that 90.2% (n = 267) slept in the peridomestic and 8.4% (n = 25) in the indoors (Table 1).

**Table 1.**
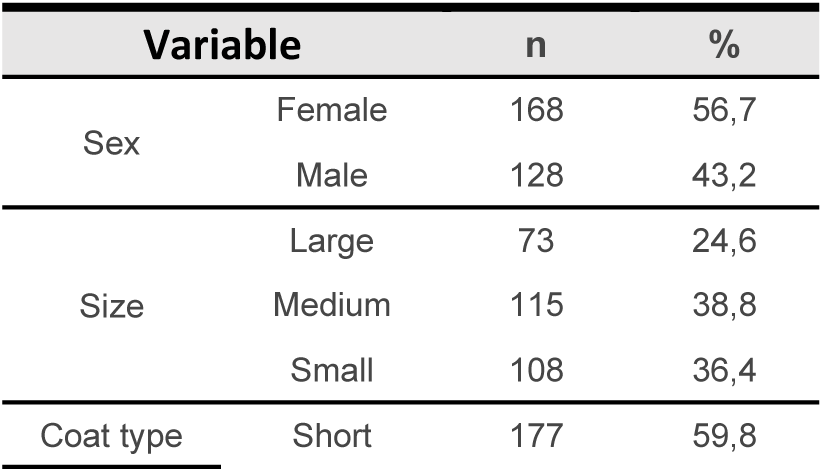
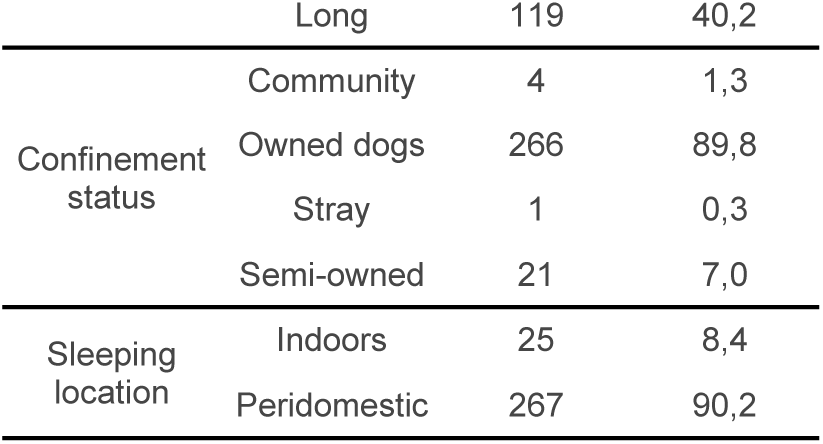
Profile of dogs included in the study, with respective absolute and relative frequencies.

### Cohort

A total of 313 dogs were enrolled in the study, with 155 allocated to the DMC+RC group and 158 to the DMC group. Subsequently, 9 dogs from the DMC+RC group and 8 from the DMC group were diagnosed with CVL and, therefore, excluded from the cohort.

During the study, high loss of follow-up was observed, reaching 37.7% (111/296) after 24 months, being higher in the DMC+RD group (43%). In the different periods evaluated, the high frequency was between baseline and T1 (25.3%), with similarity in the two groups (DMC+RD =26% and DMC=24.7%). As regards, the causes of follow-up loss in the first cycle, 20/83 (24%) were related to allergic reaction, 10/83 (12%) to change of address, 10% by refusal of the owner. Between the first and second collar cycles (T1-T2), the loss of follow-up was 24% (25/104) in the DMC+RD group while in the DMC was 0.9% (1/109) (Fig 4). Nevertheless, in the last cycle the higher loss was in the DMC group 9.6% (10/104). The median length of stay of the animals in the cohort was 532 days, being similar in the two groups (532 days for DMC+RD and 534 days for DMC).

**Fig 4.**
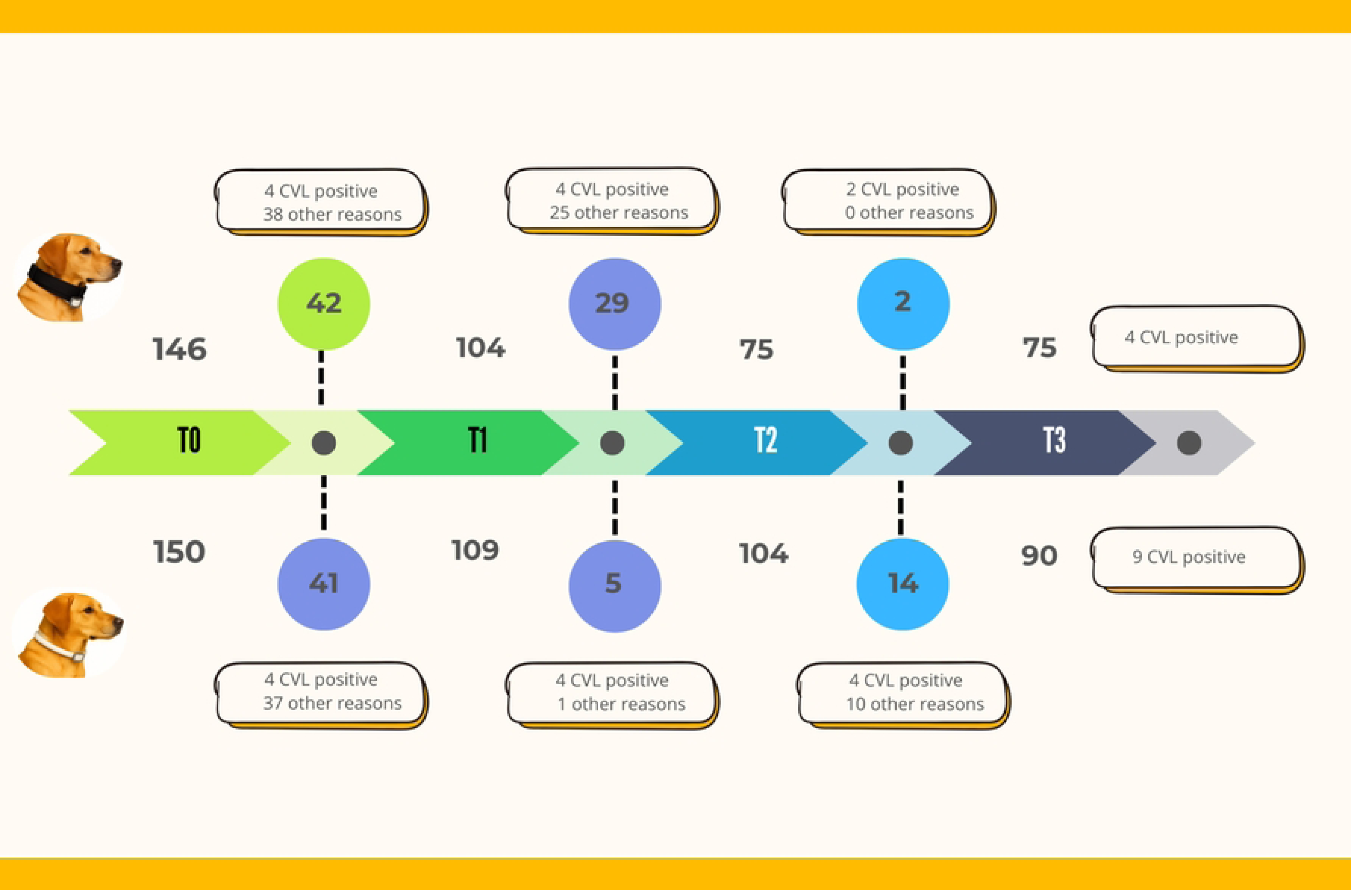
Temporal distribution of cohort participants by intervention group and follow-up period.

### Effectiveness of the collar protection

Following the initial exclusion of CVL positive animals, the adjusted baseline prevalence was 5.8% in the DMC+RD group and 5.0% in the DMC group (Fig 5). A reduction in infection prevalence was observed in both groups during the T0–T2 interval, reaching 2.6% in the DMC+RD group and 3.8% in the DMC group. However, during the T2–T3 period, prevalence increased in both cohorts, rising to 5.3% in the DMC+RC group and 10% in the DMC group (Fig 5).

**Fig 5.**
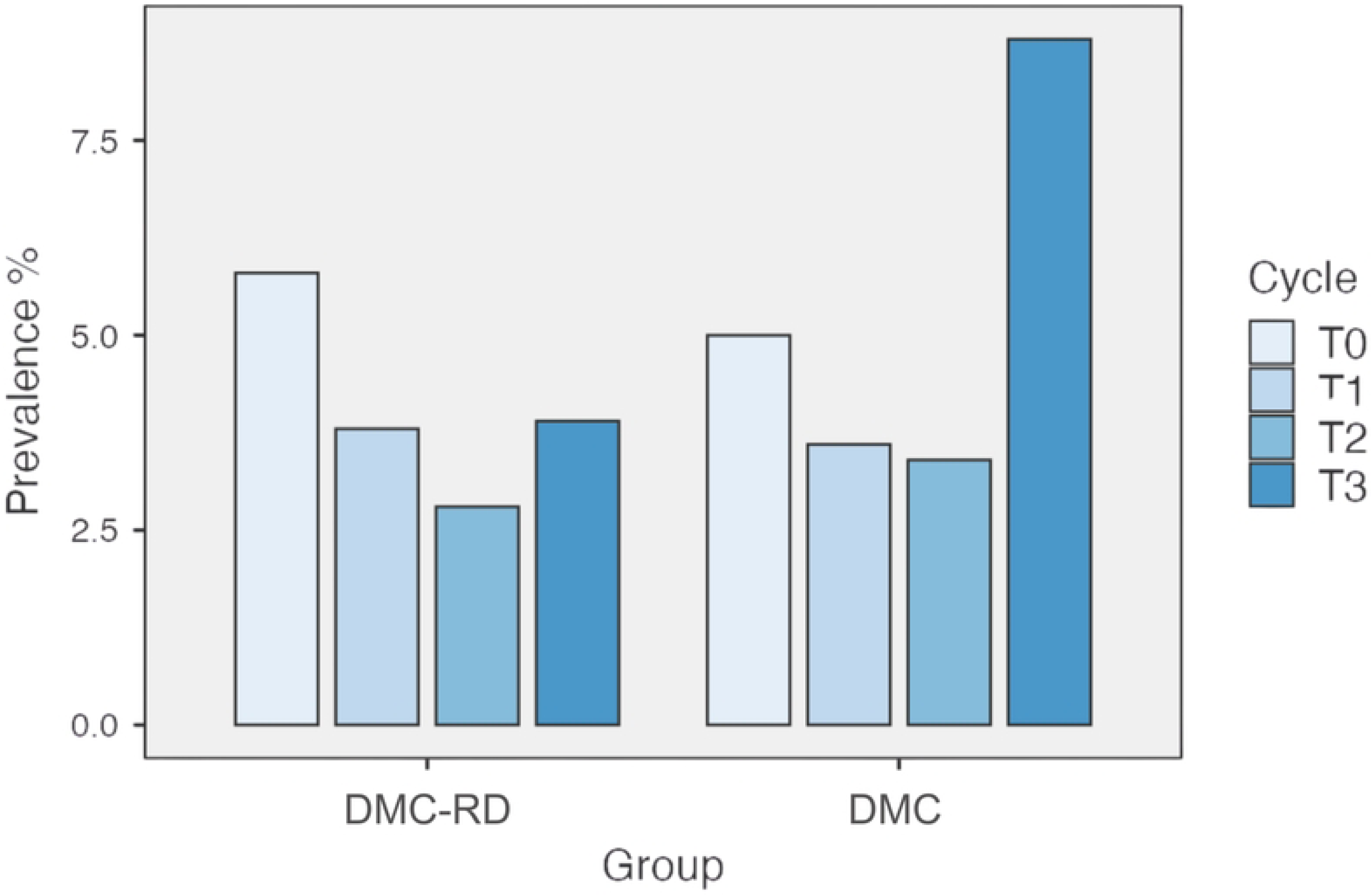
Prevalence of infection by group and observation period (T).

Regarding the chance of infection by *Le. infantum* among the groups, the results indicated that dogs in the DMC+RD group had a 41% reduced chance of infection (OR=0.59, 95% CI: 0.27–1.31), compared to the DMC.

### Effectiveness of DMC+RD in reducing canine collar loss

Comparison between the two groups as regard the duration of the collar fit in the dog showed a higher time of collar permanence in the DMC+RD. When analyzing the probability of staying with the collar over time according to the group, it was observed that dogs in the DMC+RD group had a higher probability of remaining with the collar compared to those in the DMC group (Fig 6). However, this difference was not statistically significant.

**Fig 6.**
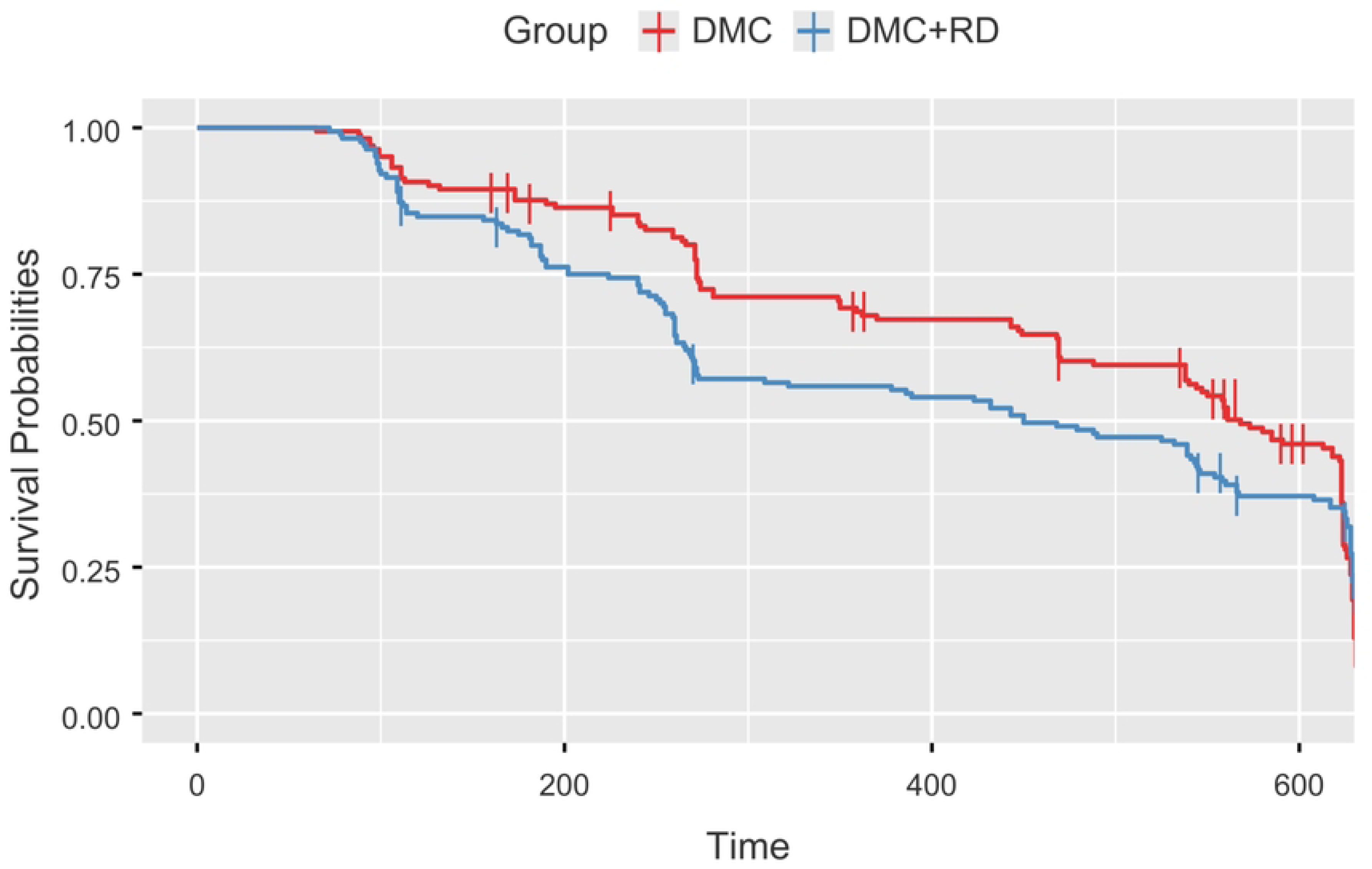
Survival curve (Kaplan-Meier) comparing the DMC group and the DMC+RD.

The association between the type of intervention and collar loss was assessed using odds ratios (OR), with estimates calculated at three distinct follow-up time points (T1, T2, and T3). Overall, there was a trend toward reduced collar loss in the group using the reinforced collar, although no statistically significant differences were observed across the evaluated time points.

At T1, the OR was 0.552 (95% CI: 0.2901 – 1.06; p = 0.0778), suggesting a possible reduction in the likelihood of collar loss in the reinforced group. At T2, the OR was 0.5948 (95% CI: 0.2916 – 1.213; p = 0.163), maintaining the pattern of lower loss occurrence, though without strong statistical evidence. At T3, the OR was 0.8769 (95% CI: 0.3513 – 2.189; p = 0.820), indicating no relevant difference between groups at this time point.

In the DMC+RD 53% of the losses occurred between 90 and 180 days during T1. At T2, 20% occurred during the first month and 27% between the second and third month. Nevertheless, 33% of the dog owners did not know exactly when the loss occurred. In T3, a high proportion of losses was observed between the first and third month.

In the DMC group 35% of the losses occurred between 150 and 180 days during T1. At T2, the time when the loss occurred was not identified by the dog owners by almost 50%. In T3, a high proportion of losses was observed in the first 90 days, representing 55% of the losses reported. At T3, similar percentages of losses were observed in the first 60 days (around 15%), and also on the 120-150 days (Fig 7).

**Fig 7.**
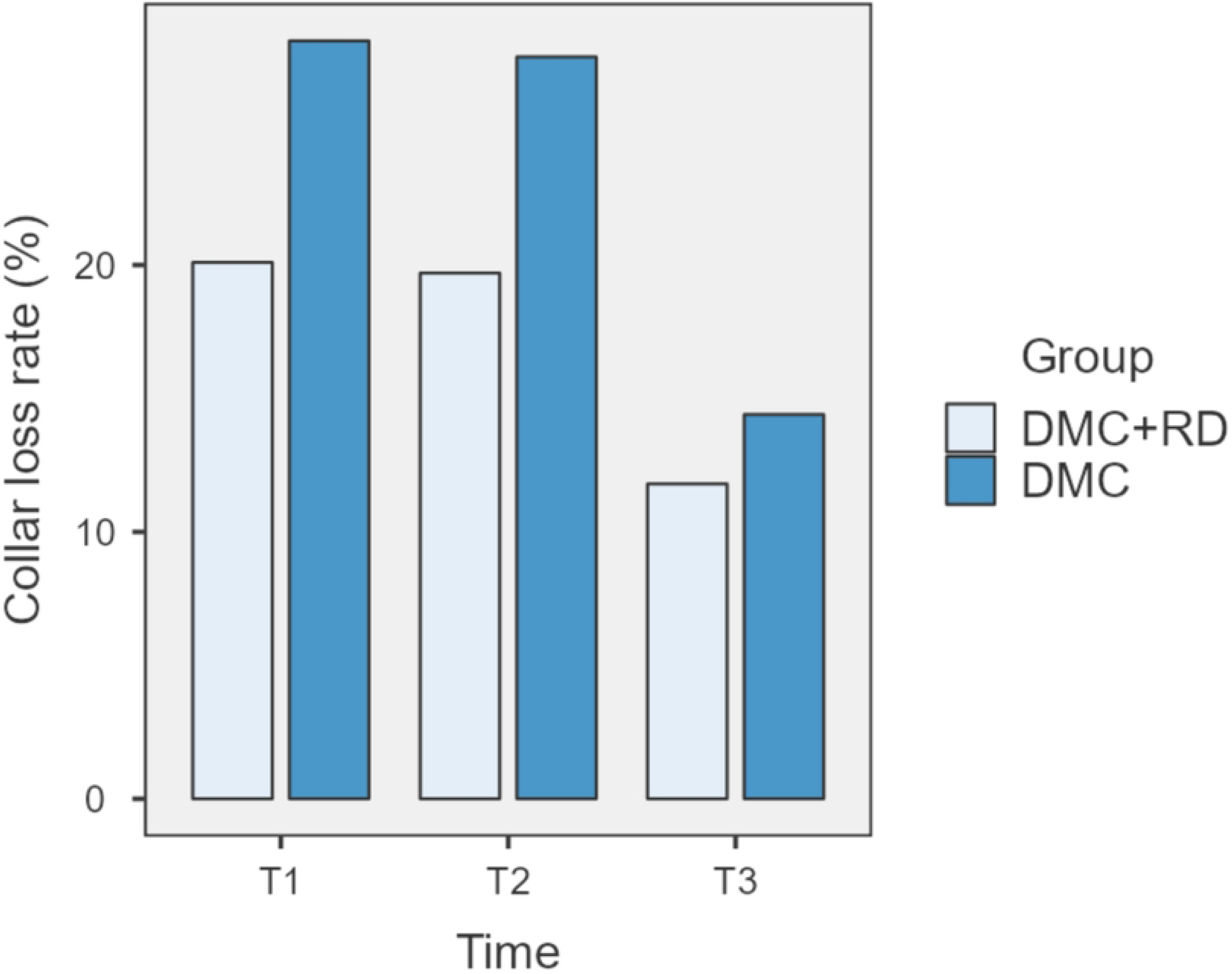
Percentage of collar losses by group and period.

## Discussion

This study describes the results of a cohort follow up of a canine population in a region under mass use of DMC for the prevention and control of VL. The sample included a predominance of females (58.1%), aged between 1 and 3 years, and a balanced distribution between small and medium-sized dogs, a profile consistent with that observed in other Brazilian endemic urban areas [22, 23, 24]. Those characteristics are common in domiciliated dogs considering ease of handling, lower space requirements, and temperament more adaptable to the domestic environment [25, 26].

In the sample a high frequency of short coats (61.1%), housed indoors (92.5%), and sleeping in the peridomicile (91.7%) was also observed. These variables are important in the VL dynamic, considering that short coats could facilitate the sandflies bites, and that sleeping outdoors (peridomicile) increases the exposure to the sandflies, and therefore their risk for CVL [27, 13]. Those results show the importance of considering those various canine population profiles, as they could influence the outcomes for the same intervention in different regions.

Regarding the cohort follow-up, a high percentage of loss of follow-up was observed, reaching almost 38% at the end of the study, being higher in the DMC+RD group (43%). These results were similar to those observed in a canine cohort in Andradina/SP when the loss of follow-up was 55%. Nevertheless, the observed losses were higher than those described by Coura-Vital [23], which reported a loss of follow-up close to 20%. Howe [28] pointed out that follow-up losses can occur due to multiple factors, including individual characteristics, risk behaviors, clinical conditions, and socioeconomic factors. In long interventions, the loss of follow up could be variable, as observed in a study conducted in Italy where it reached 35% in the first survey and 67% in the subsequent [29].

In our study, the majority of the losses occurred during the first period (T0-T1) and the main reasons for the loss of follow-up were change of address, dog donation, death of the animals by other causes, and closed houses. These results are in concordance with those reported by Coura-Vital [23], which were death, change of address, household closed, refusal, and dog escape. Other reasons such as seroconversion and euthanasia were also reported as important causes of follow-up [13]. However, its import remarks that the losses observed in our study were balanced in the two groups, and the remaining dogs in the cohort were no different from those lost [30]. This information needs to be included when multiple outcomes are evaluated to reduce potential bias

### Effectiveness of the collar protection

The effectiveness of DMC has been evaluated in different studies, showing its capacity to protect dogs from sandflies bites [11, 31, 32] and also for the reduction of human infection and canine prevalence [33, 13]. In the present study, during the first year of dog collaring (T0 - T2), a reduction in prevalence was observed, dropping from 5.8% to 2.8% in the DMC+RD group and from 5.5% to 3.4% in the DMC group, representing a reduction of 52% and 32%, respectively. Similar results were reported by Leite [17] (2018) in Camaçari (BA), where canine prevalence was reduced by 28.6% after the first intervention cycle and by another 33.3% after the second, totaling a 52.4% drop at the end of the follow-up.

However, in the last cycle of collaring (T2 -T3) there was a change of trend, increasing the prevalence in both groups, but mainly in the DMC group. Similar results were observed in a study conducted in Andradina (SP), where, after an initial drop in incidence from 10.7 to 3.9 cases per 100 dogs between T0 and T1, there was an increase to 5.3/100 dogs in T2 [13].

In a study conducted in Bahia state comparing the DMC with a control group (without), it was also observed a decrease in the prevalence of infection in both groups in the second survey, and an increase in the infection in the control group in the subsequent survey [17].

Therefore, it seems that there are other factors that affect the infection force in the endemic areas that might impact the intervention outcomes. This difference may be related to operational factors, including delayed diagnosis, failure in the removal of infected dogs, changes in the dynamics of the canine population, and the refusal of some dog owners to authorize the euthanasia of seropositive animals [13]. Another factor that could be considered is the vector population dynamic that can increase the dog-sandfly contact in different times as results of seasonal variations.

### Collar loss

The implementation of mass use of impregnated collars have different challenges including the difficulties of finding the dogs in subsequent visits to the collar fitting, closed residences, and also the loss of collars [34]. The last one can impact the protection of dogs against the sandflies bites and therefore increasing the risk of VL. In the present study a high percentage of collar loss was observed with an average of 20.4% overall. Previous studies in Brazil reported a collar loss of 41% [35]. However, it shows high variability, with low percentages observed in a study conducted in Iran, with estimated loss percentage of 1% per month [18], contrasting with another study that reports 10.3% of collar loss in that country. This variability in the collar loss could be related to the differences in the populations under intervention, operational aspects and population engagement. Among the reasons for collar loss, the dog behavior must be considered; because they can remove the collar playing or fighting; as well the collar design, as pointed out by Maroli [29], who observed that improving the collar by reinforcing the buckles contributes to the reduction of collar loss.

In this study we evaluated the impact of a reinforcement device, a Nylon collar where the DMC was attached to. A lower loss ratio was observed in the DMC+RD when compared to the DMC, however, no statistically significant differences were observed. Nevertheless, the results show that although both collars protect the *Le. infantum* infection, the odds are lower in the DMC+RD, suggesting that this device could contribute to a better result of the intervention. Further longitudinal studies with extended follow-up periods are warranted to enhance the robustness of observational findings and validate long-term outcomes

Our results also show a high proportion of dog owners who do not know when the dogs lost the collar, reaching 50% in the DMC and 33% in the DMC+RD group at T2. These results suggest low engagement of dog owners in the implementation, and low perceptions of dog health importance. It could be explained by cultural differences in the community under intervention, the risk perception and knowledge of the disease transmission, therefore, studies evaluating aspects such as acceptability, appropriateness, etc. are necessary [36]. Because the loss of collars could impact the effectiveness of the intervention [29], it is necessary to increase the population engagement by means of improving communication about the implementation, health education campaigns as well as identifying key actors to ensure the success and maintenance of the program.

Although some limitations in this study must be considered including the high loss of follow-up, it is important to highlight that they were mainly related to independent factors, also reported in previous studies [30]. Thus, our results bring important data on the application of this strategy under real conditions, promoting their large-scale use and sustainability in other regions where it could be implemented, and also the control program improvement.

High effectiveness of DMC applied in a mass population control program was observed, as well that the improvement of the collar design could contribute to reducing the loss of collars and increase the protection. Nevertheless, other factors related to the owners’ engagement, operational difficulties, and dog population dynamic, can influence the outcomes of the implementation strategy. Future studies are necessary to evaluate the impact of programmatic and educational initiatives to promote community adherence and sustainability.

## Data Availability

All relevant data are within the manuscript and its Supporting Information files.

## Acknowledgments

We would like to thank to the dog owner that accept to participate of the study, to the entire team of endemic disease control agents from the municipality of Montes Claros, especially to Ronaldo Cardoso dos Santos, Marcelo Dias Soares, Douglas Aparecido de Oliveira, and José Renilson Ramos da Cruz, for their logistical support during the blood sampling of animals.

